# Sex differences in the risk of cataract associated with type 2 diabetes: a Mendelian randomization study

**DOI:** 10.1101/2021.08.05.21261684

**Authors:** Haoyang Zhang, Xuehao Xiu, Yuedong Yang, Yuanhao Yang, Huiying Zhao

## Abstract

Type 2 diabetes (T2D) is a recognized risk factor for developing cataract. However, it is unclear if the shared genetic variance and potential genetic causal relationship between T2D and cataract are different for males and females. We evaluated sex-specific genetic correlation (*r*_g_) and putative genetic causality between the two diseases by using linkage disequilibrium score regression (LDSC) and six Mendelian randomization (MR) approaches after leveraging large-scale population-based genome-wide association studies (GWAS) summary of T2D and cataract. Application of LDSC found a significant genetic correlation between T2D and cataract in East Asian males (*r*_g_=0.68, 95% confident interval [CI]=0.17 to 1, p-value=8.60×10^−3^) but a non-significant genetic correlation in East Asian females (*r*_g_=0.25, CI= -0.02 to 0.52, p-value=8.38×10^−2^). MR analyses indicated a consistently stronger (paired t-test |t|=5.87, p-value=2.04×10^−3^) causal effect of T2D on cataract in East Asian males (liability OR=1.20 to 1.41, p-value=5.86×10^−27^ to 6.60×10^−6^) than in females (liability OR=1.12 to 1.21, p-value=2.02×10^−14^ to 1.82×10^−2^). In Europeans, the LDSC analysis suggested a close significant genetic correlation between the two diseases in males (*r*_g_=0.20, 95% confident interval [CI]=0.08 to 0.32, p-value=7.00×10^−4^) and females (*r*_g_=0.17, CI= 0.05 to 0.29, p-value=4.90×10^−3^); but the MR analyses provided weak evidences on a causal relationship between the two diseases in both sexes. These results presented the first evidence on sex difference of the casual relationship between cataract and T2D in East Asians, and supported a potential genetic heterogeneity of the shared genetics underlying T2D and cataract between East Asians and Europeans in both sexes.

## Introduction

Type 2 diabetes (T2D) is one of the most prevalent chronic diseases(1) and a recognized risk factor for cataract development(2). Previous studies have revealed both phenotypic(2, 3) and genotypic(4, 5) associations between the two diseases. Although a higher risk of cataract in female than in male was well-established by multiple phenotypic studies(6-8), the sex difference of cataract risk in T2D patients was still inconclusive and being studied. Olafsdottir et al.(9) observed the risk of nuclear cataract in T2D females was higher than that of T2D males from an Icelandic cohort (*N*=531). Another Indian T2D cohort study (*N*=779) conducted by Srinivasan et al. also identified an increased risk of posterior subcapsular cataract or nuclear cataract in females compared to males. However, a recent study(10) in Australia, Fremantle Diabetes Study Phase II (FDS2) (*N*=6316), suggested sex was not a risk factor for incidence of intraocular lens implantation. Similarly, another Taiwan cohort study of 578 T2D participants also found that the sex difference of cataract risk did not exist in the multivariate analysis (controlling covariates like Body mass index [BMI], Systolic blood pressure [SBP], and Hemoglobin A1c[HbA1c])(11). These results suggested that potential confounding factors and limit sample size may influence the estimation of sex difference in cataract risk. Thus, it is worthwhile to investigate such a sex-differential effect on the basis of genetic correlation and causal relationships by leveraging large population-based genetics studies.

With the development of genome-wide association studies (GWAS) during past decades, multiple methods were exploited to investigate the shared genetic architecture and genetic casual relationships between traits. For example, by examining the linkage disequilibrium (LD) score from single nucleotide polymorphisms (SNPs) across two traits, linkage disequilibrium score regression (LDSC)(12) was designed to quantify the genetic correlation feasibly by measuring the correlation of LD scores from two traits. For traits with significant genetic correlation, Mendelian randomization (MR)(13) is a popular tool for searching potential causal effects from a trait (i.e., exposure) to another (i.e., outcome) by using independent SNPs strongly associated with the exposure trait as instruments. MR exploits the concept of natural random allocation of genetic variant to mimic the randomization procedure in randomized controlled trials (RCTs). The genetic causality inferred by MR is less-biased because the allocation of genetic variant is generally not correlated with both exposure and outcome(13). A special application of MR is summary-data-based Mendelian randomization (SMR)(14) that treats gene expression in expression quantitative trait loci (eQTL) summary data as “exposure” and a focal trait as “outcome”. Identifying potential functional genes related with a genetic correlation, SMR may provide new insights for the understanding of shared genetic etiology (15). Application of MR analysis indicated a potential causal effect of T2D on cataract in East Asians(15), but it is far from clear if a sex difference exists in the causal relationship between T2D and cataract.

In this study, we utilized sex-stratified GWAS summary statistics of T2D and cataract from multiple sources (i.e., Asian Genetic Epidemiology Network [AGEN](16), BioBank Japan [BBJ](17), and UK Biobank (UKB)(18)) to investigate the genetic relationship between T2D and cataract in both East Asians and Europeans. We performed sex-stratified LDSC and MR analysis to measure the genetic correlation and genetic causality between T2D and cataract, and tested if the genetic correlation and genetic causality were different between males and females. Then, we implemented SMR analysis (14) to identify potential functional genes underlining the putative sex-differential causality. The flowchart of our study was shown in Fig. 1.

**Figure 1.**
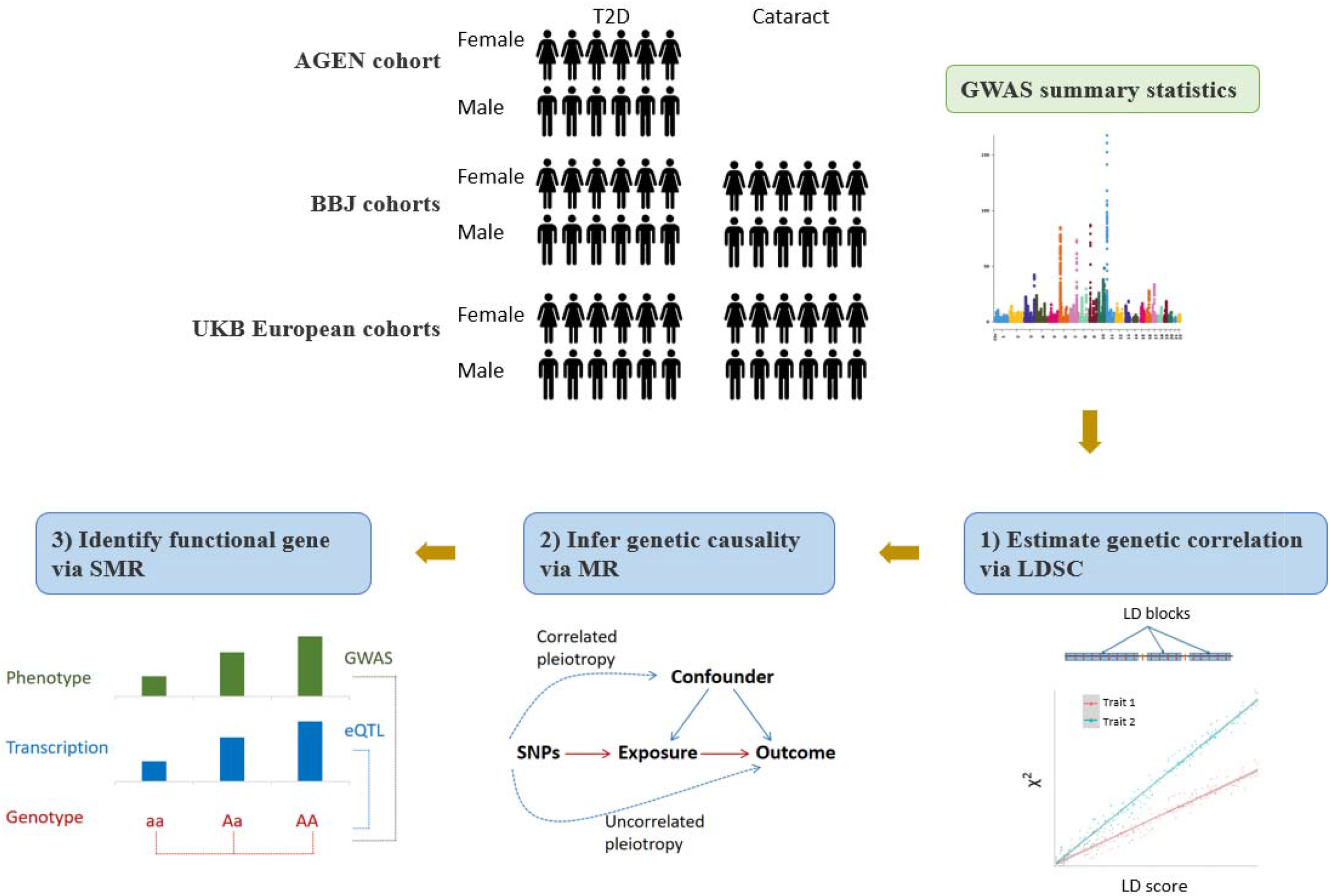
Outline of analysis performed in the study. Utilizing large-scale population-based GWAS summary statistics from multiple source, the study conducted a series of sex-stratified analysis to investigate the shared genetics underling T2D and cataract. It included three sections: 1. Measuring the sex-stratified genetic correlation between T2D and cataract with LDSC; 2. Inferring the sex-stratified causal relationship between T2D and cataract using six MR approaches; 3. Identifying candidate functional genes underlying the putative sex-differential causality between T2D and cataract with SMR. LDSC: linkage disequilibrium score regression; MR: Mendelian randomization. SMR: summary data-based Mendelian randomization.

## Methods and materials

### Datasets

We downloaded the sex-stratified hg19-based GWAS summary statistics of T2D (*N*_case_=40,250 [male=25,705, female=14,545], *N*_control_=170,615 [male=82,774, female =87,841])(19) and cataract (*N*_case_=24,622 [male=11,641, female=12,981], *N*_control_=187,831 [male=97,706, female=90,125])(19) from the BBJ (http://jenger.riken.jp/en/), namely BBJ-T2D GWAS and BBJ-cataract GWAS. BBJ(17) was a large database including phenotype and genotype data from around 200,000 Japanese individuals (proportion of male=53.10%; average baseline age at 62.70 for men and 61.50 for women). In BBJ, both T2D and cataract were diagnosed by physicians; the sex-stratified GWAS of two diseases were performed using a linear mixed model via SAIGE (20) adjusted by age and top five genetic principal components.

We downloaded another sex-stratify T2D GWAS summary statistics from AGEN (https://blog.nus.edu.sg/agen/summary-statistics/t2d-2020/), namely AGEN-T2D GWAS. The AGEN-T2D GWAS (16) was generated using a fixed-effect inverse-variance-weighted meta-analysis via METAL(21) from 22 East Asian population-based cohorts (*N*_case_=117,339 [male=28,027, female=89,312], *N*_control_=162,425 [male=27,370, female=135,055]). The East Asian samples of AGEN-T2D GWAS was approximately independent to that of BBJ-T2D GWAS. Thus, we employed AGEN-T2D and BBJ-cataract to investigate the genetic correlation, as well as the causal relationship between two diseases in East Asians. Parallel analysis between BBJ-T2D and BBJ-cataract was also performed to investigate the robustness of the findings.

### Genome-wide association study on T2D and cataract of Europeans from UK Biobank

We generated the sex-stratified GWAS summary statistics of T2D (*N*_case_=29,267 [male=17,688, female=11,579], *N*_control_=473,237 [male=211,434, female= 261,803]) and cataract (*N*_case_=33,842 [male=14,758, female=19,084], *N*_control_=468,662 [male=214,364, female=254,298]) using the European cohorts in UK Biobank (18), namely UKB-T2D GWAS and UKB-cataract GWAS. UK Biobank is a prospective study with over 500,000 participants across different population (proportion of male=45.60%; average baseline age at 56.74 for men and 56.35 for women). We determined the case of T2D and cataract patients by ICD-10 code (UKB field 41270). An individual with ICD-10 code of E11 (non-insulin-dependent diabetes mellitus) was collected as a T2D patient while an individual with ICD-10 codes of H25 (senile cataract) or H26 (other cataract) was collected as a cataract patient. The genetic location of the genotypic data was based on hg19. The sex-stratified GWAS was performed by BOLT-LMM (22) adjusting for age, sex, and cryptic relatedness.

### Estimation of heritability and genetic correlation

We performed sex-stratified LDSC(12, 23) to estimate the single-trait liability-scale heritability (*h*^2^) of T2D and cataract, and the cross-trait genetic correlation (*r*_g_) between: 1. AGEN-T2D and BBJ-cataract, 2. BBJ-T2D and BBJ-cataract, and 3. UKB-T2D and UKB-cataract.

We used the genotype data of 481 East Asians and 489 Europeans in the 1000 Genomes as reference genome. The reference genome and the pre-calculated LD scores were downloaded from the LDSC website (https://alkesgroup.broadinstitute.org/LDSCORE/). We conducted quality control for SNPs inputting into LDSC analysis by: 1. filtering SNPs using the HapMap3(24); 2. removing the SNPs if they were strand-ambiguous; 3. removing SNPs whose minor allele frequency (MAF)<0.01; 4. removing SNPs located in the major histocompatibility complex (MHC) region (chromosome 6: 28,477,797–33,448,354)(25).

Single-trait LDSC was performed to estimate the sex-stratified liability-scale *h*^*2*^ of T2D and cataract. For East Asians, we set the population prevalence of T2D (male: 5.90%(26), female: 7.00%(26)) and cataract (male: 40.60%(27), female: 42.33%(27)) according to cohort studies conducted by Wang et al.(26) and Tang et al.(27). For Europeans, we set the population prevalence of T2D (male: 3.80%(28); female: 3.10%(28)) and cataract (male: 30.02%(29), female: 40.69%(29)) according to a health survey conducted by National Health Service (NHS) Health and Social Care Information Centre(28) and a cross-sectional study conducted by Reidy et al.(29).

Cross-trait LDSC was applied to measure the sex-stratified genetic correlation between T2D and cataract. We performed cross-trait LDSC twice, with and without a constrained intercept, since constraining the intercept may increase the accuracy of estimation when the impact from population stratification is limited. A significant *r*_g_ was determined with p-value<0.05/2=0.025. The differences of *r*_g_ between two sexes were evaluated by Z test.

### MR analyses for genetic causality

The sex-stratified causal relationship between T2D and cataract was evaluated by multiple MR approaches. Inverse-variance-weighted (IVW) method(30) is a basic MR method that integrates the GWAS effect ratios of instrumental SNPs between exposure and outcome in a fixed effect meta-analysis model. However, the horizontal pleiotropy(13) (i.e., a potential confounding that instrumental SNPs effect on outcome through non-causal pathways) may affect measurement of casual relationship between traits for IVW. Horizontal pleiotropy contains uncorrelated pleiotropy and correlated pleiotropy. Uncorrelated pleiotropy is a type of pleiotropy whose effect on outcome is independent to exposure, and correlated pleiotropy is another type of pleiotropy whose effect on outcome is related with exposure(13). To reduce the probability of false positive in MR analysis, expect for IVW, five other MR methods(i.e., MR-Egger (31), generalized summary-data-based Mendelian randomization [GSMR] (32), weighed median (33), weighted mode (34), and the causal analysis using summary effect estimates [CAUSE] (35)) with different assumptions on horizontal pleiotropy were also employed(36).

Comparing to IVW, MR-Egger made an improvement to account for uncorrelated pleiotropy by adding an intercept in the IVW regression model. In GSMR analysis, SNPs were excluded if they involve in uncorrelated pleiotropy by a heterogeneity in dependent instrument (HEIDI) outlier test. The weighted median model measures the causal effect as the weighted median of GWAS estimates ratios to reduce the impact from pleiotropic SNPs. The weighted mode model loosens the assumption of weighted median model (i.e., pleiotropy is existed in less than half of instrumental SNPs), and measures the causal effect from the most frequent (mode) instrumental SNPs. For these five methods (IVW, MR-Egger, GSMR, weighted median, and weight mode), we selected independent instrumental SNPs with GWAS p-value<5×10^−8^, and clumped with LD *r*^2^<0.05 within 1000-kb window using PLINK 1.9(37) according to the same 1000 Genomes reference described above(38). CAUSE (35) is a more powerful method to identify the causality from both types of pleiotropy by pruning SNPs with a GWAS p-value threshold at 1×10^−3^ and a LD *r*^2^ threshold at 0.1. It also conducted a model comparison between the causal model (i.e., instrumental SNPs act on exposure and outcome through both causal pathways and shared factors) and sharing model (i.e., instrumental SNPs act on exposure and outcome only through shared factors) based on the expected log pointwise posterior density (ELPD) test.

We used six MR analysis approaches to examine the genetic causality between T2D and cataract. The MR analyses were performed by R packages “cause”, “gsmr” and “TwoSampleMR”. Instrumental SNPs with MAF>0.01 or located within the MHC region were excluded(22). A significant causal relationship was determined if the MR p-value reached Bonferroni-corrected level (i.e., less than 0.05/12≈4.17×10^−3^, adjusted by six bi-directional tests). The logit-scale MR estimates (i.e., *β*_MR_) were further transformed to liability-scale according to the formula proposed by Byrne et al^(39)^:

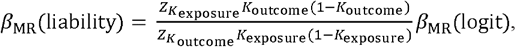

where *K*_exposure_ and *K*_outcome_ represent the population prevalence of exposure and outcome while 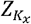 and 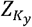 represent the value of standard normal distribution at such prevalence. We assumed the population prevalence is the same as that used in estimating the liability heritability. The liability *β*_MR_ was then converted into odds ratio (OR). The sex difference of causal effect between two sexes estimated with liability *β*_MR_ was evaluated by paired t-test.

### SMR analysis to identify candidate functional genes underling the genetic relationship of T2D and cataract

If a sex-differential genetic relationship was suggested by MR analysis, we further explored potential functional genes associated with the sex-differential causal relationship via SMR(14). SMR can detect functional genes associated with a trait by treating gene expression as “exposure” and a trait as an “outcome” (e.g., T2D) with top cis-eQTL (GWAS p-value<5×10^−8^) associated with each gene (19,250 probes, assessed from a blood-based cis-eQTL summary dataset(40), URL: https://eqtlgen.org/cis-eqtls.html). We applied SMR analysis on each single-trait GWAS, as well as the cross-trait GWAS of T2D and cataract generated with inverse-variance-weighted meta-analysis via MTAG(41).

To exclude associations introduced by linkage (e.g., a top SNP was in LD with causal SNPs which affecting a gene expression and the trait, respectively), we conducted HEIDI test on each SMR estimates. Significant functional genes were determined with SMR p-value<0.05/19250≈2.60×10^−6^ and HEIDI p-value>0.05 from at least 10 SNPs.

## Results

### The genetic correlation between T2D and cataract in males and females

Single-trait LDSC indicated the liability-scale heritability of T2D and cataract in males (AGEN-T2D: *h*^2^=0.22, p-value=6.94×10^−20^; BBJ-cataract: *h*^2^=0.02, p-value=1.51×10^−1^) and in females (AGEN-T2D: *h*^2^=0.32, p-value=2.15×10^−11^; BBJ-cataract: *h*^2^=0.02, p-value=1.45×10^−1^) were approximately equal in East Asians. Using cross-trait LDSC, we identified a significant genetic correlation between AGEN-T2D and BBJ-cataract in males (*r*_g_ =0.68, 95% confident interval [CI]=0.17 to 1, p-value=8.60×10^−3^) but not in females (*r*_g_=0.25, CI= -0.02 to 0.52, p-value=8.38×10^−2^). The genetic correlation in females were lower than that in males although the gap was non-significant (|Z|=1.68, p-value=9.30×10^−2^). The analysis using BBJ-T2D GWAS and BBJ-cataract estimated a higher genetic correlation (males: *r*_g_=0.72, 95%CI=0.23 to 1, p-value=4.10×10^−3^; females: *r*_g_=0.45, 95%CI=0.08 to 0.82, p-value=1.57×10^−2^) than using AGEN-T2D and BBJ-cataract, suggesting a potential inflation introduced by partial sample overlap from two BBJ cohorts.

To estimate the sex-stratified genetic correlation between T2D and cataract in Europeans, we carried out GWAS analysis based on the European cohorts in UK Biobank. The Manhattan and quantile–quantile plots of the distribution of GWAS p-value were displayed in Fig. S1 and Fig. S2. The effect size of genome-wide significant SNPs with GWAS p-value<5×10^−8^ were detailed in Table S1. Single-trait LDSC detected a mild inflation of our GWAS analysis (UKB-T2D male: λgc=1.20, LDSC intercept=1.03; UKB-T2D female: λgc=1.25, LDSC intercept=1.04; UKB-cataract male: λgc=1.10, LDSC intercept=1.01; UKB-cataract female: λgc=1.09, LDSC intercept=1.02). The liability-scale heritability of cataract in males (*h*^2^=0.15, p-value=7.03×10^−15^) is close to the heritability of cataract in females (*h*^2^=0.11, p-value=4.80×10^−9^) while that of T2D in males (*h*^2^=0.15, p-value=6.03×10^−45^) was lower than that in females (*h*^2^=0.29, p-value=5.16×10^−52^). The genetic correlation between the two diseases in males (*r*_g_=0.20, p-value=7.00×10^−4^) was approximately equal (|Z|=0.33, p-value=0.74) to that in females (*r*_g_=0.17, p-value=4.90×10^−3^). The detailed statistics of the LDSC analysis was listed in Table S2.

### Sex differences in the putative causality of T2D on cataract

Based on the strong genetic correlation between T2D and cataract in East Asians, we implemented multiple MR methods to infer the sex-specific causal relationship between the two diseases (see Fig. 2 and Table. S3). By analyzing AGEN-T2D GWAS and BBJ-cataract GWAS, we identified robust evidences for a putative causal effect of T2D on cataract for East Asian males (liability OR=1.20 to 1.41, p-value=5.86×10^−27^ to 6.60×10^−6^), with all of six MR models surpassing the Bonferroni-corrected significance threshold (p-value<4.17×10^−3^) (Fig. 2a). We also observed suggestive evidence for the putative causal effect of T2D on cataract in females (liability OR=1.12 to 1.21, p-value=2.02×10^−14^ to 1.82×10^−2^) with four of six MR models (except for MR-Egger and weighted mode model) reaching the Bonferroni-corrected significance threshold. Notably, the estimated MR odds rations (ORs) for males was significantly stronger than that for females (paired t-test |t|=5.87, p-value=2.04×10^−3^). The results of CAUSE suggested that comparing to healthy individuals, a male with T2D had approximately 1.20 times risks for cataract, and the risk would be reduced to 1.12 for females (Fig. 3a). The sex difference in causal effect from T2D to cataract was also supported by a parallel analysis between BBJ-T2D and BBJ-cataract (paired t-test |t|=20.45, p-value=5.18×10^−6^) (Fig. 2b & Fig. 3b). In the reverse direction, there was no evidence for a causal effect of cataract to T2D in both sexes (male: liability OR = 1.01 to 1.24, p-value=1.97×10^−1^ to 9.37×10^−1^; female: liability OR = 0.97 to 1.09, p-value=5.20×10^−1^ to 9.58×10^−1^) (Fig. 2c).

**Figure 2.**
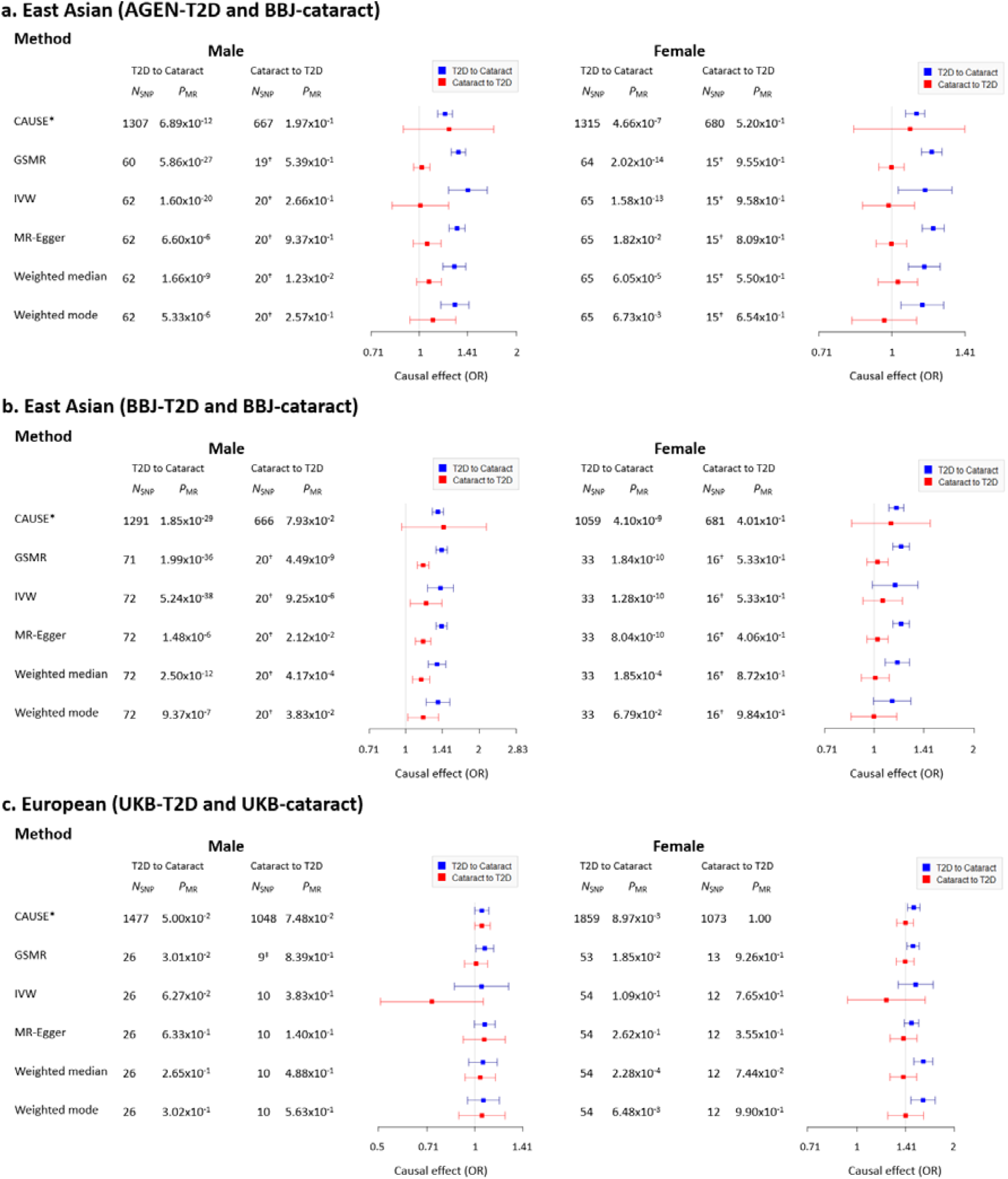
a. The sex-stratified bi-directional causal effect between T2D and cataract in East Asians estimated by six MR methods using AGEN-T2D and BBJ-cataract. b. The sex-stratified bi-directional causal effect between T2D and cataract in East Asians estimated by six MR methods using BBJ-T2D and BBJ-cataract. c. The sex-stratified bi-directional causal effect between T2D and cataract in European estimated by six MR methods using UKB-T2D and BBJ-cataract. In the forest plot, the blue bar represents the MR estimates at direction of T2D to cataract (i.e., T2D as exposure and cataract as outcome), the red bar represents the MR estimates at direction of cataract to T2D (i.e., cataract as exposure and T2D as outcome). CAUSE: causal analysis using summary effect estimates; GSMR: generalized summary-data-based Mendelian randomization; IVW: inverse variance weighted. ^*^CAUSE recruited independent instrumental SNPs with GWAS p-value <1×10^−3. **†**^IVW, MR-Egger, GSMR, weighted median, and weighted mode recruited ‘proxy’ independent instrumental SNPs with GWAS p-value <1×10^−5. **‡**^10 instrumental SNPs were provided for GSMR analysis, and 1 SNP was removed by HEIDI test.

**Figure 3.**
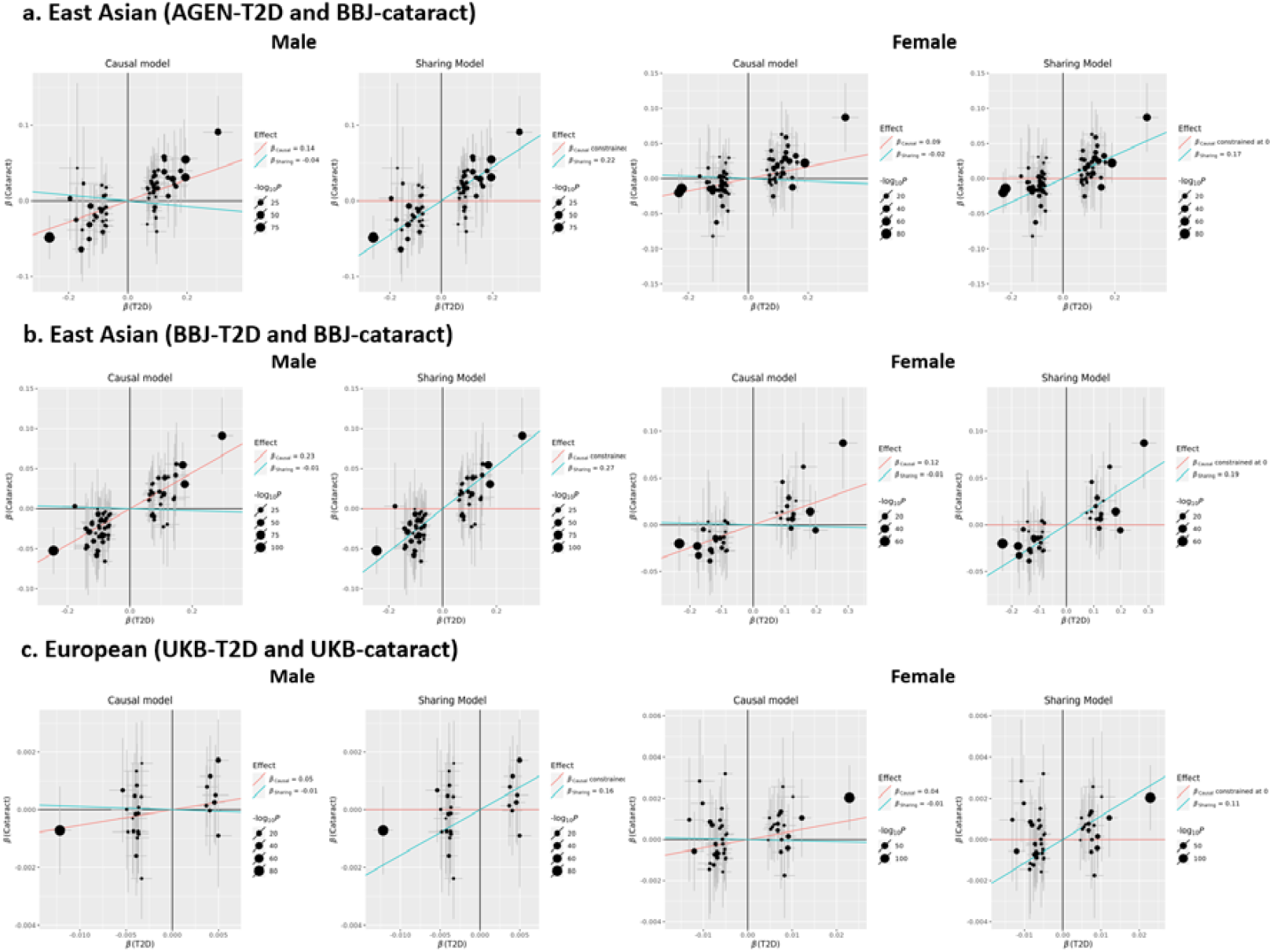
The sex-stratified CAUSE fitted causal model and sharing model using T2D as exposure and cataract as outcome. a. CAUSE models in East Asians using GWAS summary of AGEN-T2D and BBJ-cataract. b. CAUSE models in East Asians using GWAS summary of BBJ-T2D and BBJ-cataract. c. CAUSE models in Europeans using GWAS summary of UKB-T2D and UKB-cataract. Points represent instrumental SNPs with GWAS p-value<5×10^−8^ and their size represent the magnitude of GWAS p-value. Error bars stand for the 95% confidence intervals (CIs) of the SNP effect sizes based on the GWAS of T2D (x-axis) and cataract (y-axis). Blue line represents the estimated sharing effect between T2D and cataract. Red line represents the estimated causal effect of T2D on cataract, which would be constrained at zero in the sharing model. CAUSE: causal analysis using summary effect estimates.

These MR results from T2D to cataract were less likely to be influenced by the horizontal pleiotropy (Table S4) because of the non-significant and close-to-zero MR-Egger intercept (−0.008 for male and 0.004 for female, both p-value>0.05) and the better fitness of causal model than sharing model in CAUSE (ELPD p-value<0.05 in both sexes).

When the MR analysis was applied in Europeans, we found the causal effect of T2D on cataract was non-significant (male: liability OR=1.05 to 1.07, p-value=3.01×10^−2^ to 6.33×10^−1^; female: liability OR=1.04 to 1.13, p-value=8.97×10^−3^ to 2.62×10^−1^) in both males and females, as well as in the reverse direction (Fig. 2c). As revealed by CAUSE, comparing to the sharing model, the better fitness of causal model in Europeans was less significant (ELPD p-value=1.94×10^−1^ for males and 7.14×10^−2^ for females) than that in East Asians (ELPD p-value=1.33×10^−2^ for males and 1.31×10^−2^ for females), suggesting a higher pleiotropy in the analysis of Europeans than in East Asians (Fig. 3 & Table S4).

### Candidate genes likely to be involved in the shared genetics between T2D on cataract in East Asian

To identify functional genes associated with the sex difference on the risk of cataract in T2D patients, we applied SMR to the cross-trait meta-analysis GWAS (i.e., T2D and cataract) as well as the single-trait GWAS. The cross-trait meta-analysis identified five genes (*TLE1, BRD3, AP3S2, KCNJ11*, and *KLHL42*) associated with the cross-trait of T2D and cataract in East Asian males (Table 1). Among them, two genes, *BRD3* (*β*_SMR_=-0.14±0.03, SMR p-value=9.28×10^−7^, HEIDI p-value=0.20 from 14 SNPs) and *KLHL42* (*β*_SMR_=0.15±0.03, SMR p-value=1.56×10^−6^, HEIDI p-value=0.06 from 20 SNPs) were not identified to be associated with individual traits by SMR analysis. These two genes may play important roles in causing cataract in males of East Asian with T2D. When analyzing on East Asian females, we only identified one gene, *KCNJ11* (*β*_SMR_=0.38±0.07, SMR p-value=1.15×10^−8^, HEIDI p-value=0.11 from 16 SNPs) as associated with the cross-trait of T2D and cataract. Interestingly, *KCNJ11* was also identified as a functional gene in both the single trait analysis of T2D and the cross-trait analysis of East Asian males.

**Table 1.**
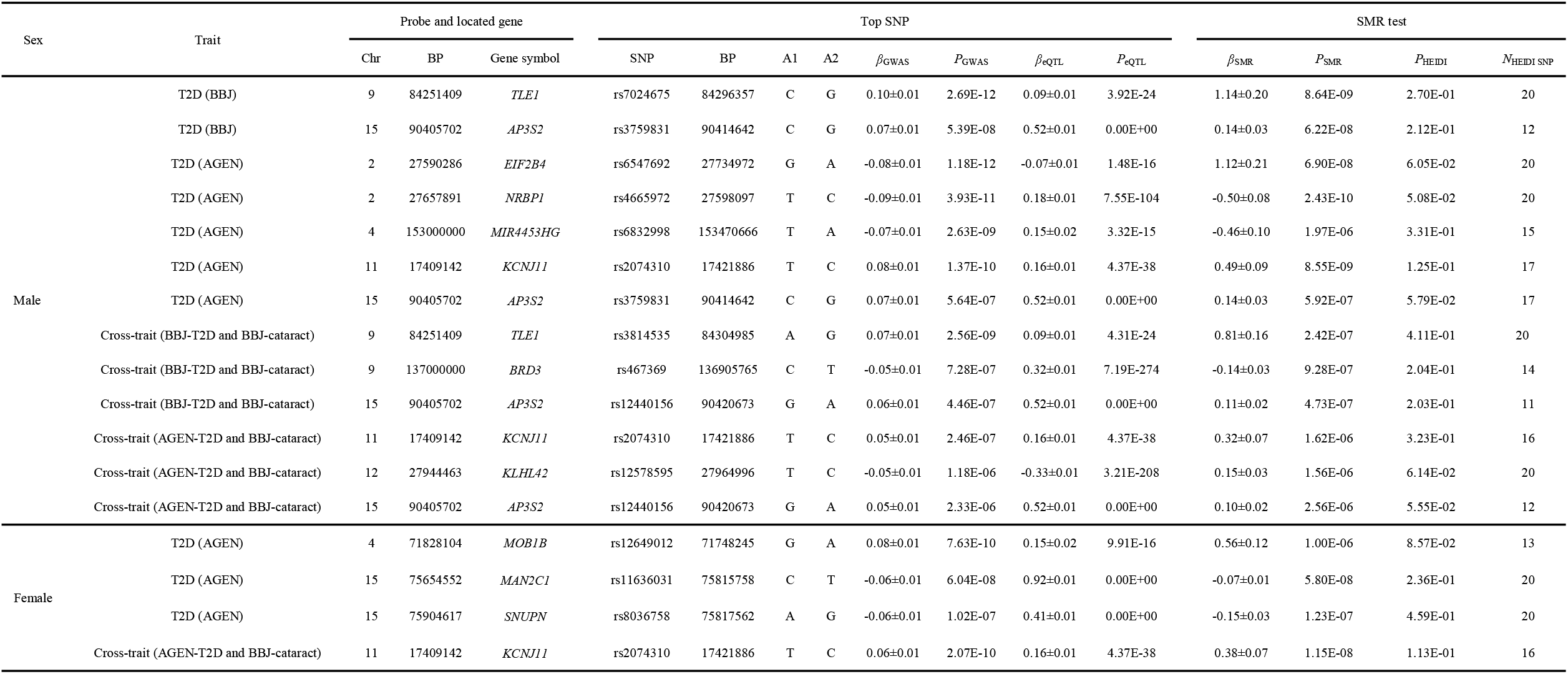
Candidate genes involving in the shared genetics between T2D and cataract in East Asians.

## Discussion

Previous epidemiology studies have paid a lot of efforts to explore the sex difference on pathways associated with co-occurrence of T2D and cataract by cross-sectional investigations and cohort studies (9, 11, 42). However, these studies are potentially limited due to their small sample size of patients co-occurring both diseases and laborious covariates collection process. Importantly, the knowledge of sex difference on the basis of shared genetics underling two diseases was limited. In the study, by using the large population-based sex-stratified GWAS summary, we performed LDSC and MR analysis to investigate the sex-differential effect in genetic correlation and causal relationship between T2D and cataract. We found that T2D patients in East Asian males have probably higher risk in developing cataract than females, and such a sex-differential casual effect was not observed in Europeans. To our knowledge, this is the first attempt to explore the sex difference in potential genetic causality between T2D and cataract using population-based GWAS summary statistics.

Application of six MR methods consistently disclosed that T2D has higher putative causal effect on cataract in East Asian males than in females. Remarkably, the sex difference in the causal relationship between T2D and cataract were incomparable between East Asians and Europeans. The potential heterogeneity of the shared genetics between two diseases in East Asians and Europeans was investigated by LDSC analysis (Table S5). In results, we observed a significant genetic correlation of T2D between two populations for both sexes, with estimated *r*_g_ in the range of 0.67 to 0.71. However, LDSC provided weak evidence (p-value>0.1) for a cross-population genetic correlation on cataract. These results suggested the cross-population heterogeneity of genetic correlations, as well as causal relationships, may be mostly due to the heterogeneity of cataract.

The previous epidemiology studies have widely accepted that the females have higher risk to suffer from cataract than males (6-8, 10, 43-46) while a number of studies suggested that sex was not a risk factor for cataract in people with T2D (10, 11, 47-49). A possible explanation to this difference is that some male-specific covariates may have a role in the development of cataract via intersecting with T2D. Moreover, our study further revealed a stronger putative casual effect of T2D on cataract in males than females in East Asians, suggesting the consequence of the interaction between male-specific covariates and T2D on cataract development may be not merely an equal risk of cataract. Further investigations are required to clarify such sex-specific factors in relationship between T2D and cataract.

The SMR analysis on the cross-trait of T2D and cataract revealed two male-specific functional genes, *KLHL42* and *BRD3*, to be relevant to the genetic causality of T2D on cataract in East Asian males. *KLHL42* encoded a protein in Kelch-like family (kelch like family member 42), and it was known to be a susceptible gene of T2D (50, 51). *BRD3* was reported to be associated with Brain-derived neurotrophic factor (BDNF) by a GAWS on Europeans (52). BDNF was revealed to be associated with increased risks of various diseases including T2D(52). The SMR analysis on East Asian males identified *KCNJ11* as associated with T2D while the SMR analysis on the cross-trait of T2D and cataract of East Asian females also revealed *KCNJ11* as a female-specific functional gene relevant to the genetic causality of T2D on cataract. *KCNJ11* is involved in the encoding of potassium channel and is reported to be associated with both T2D and congenital cataract in GWAS study of East Asians(53, 54). This study offered us the evidence that *KCNJ11* may be a candidate gene associated with putative casual effect of T2D on cataract in East Asian males, but its role in females is still unclear.

Our study has two limitations. First, since there were no other published European sex-stratified T2D and cataract GWAS apart from that in UKB, we conducted analysis between two GWAS summary based on the sample cohort. Such an operation may introduce a sample-overlap bias in our European analysis. Secondly, we used a loosen p-value threshold, 1×10^−5^ to obtain at least 10 instruments in testing the causal effect of cataract on T2D in East Asians in MR analysis. This operation may violate the assumption of GSMR (GSMR recommended at least 10 independent SNPs with GWAS p-value <5×10^−8^ associated with exposure to maintain the study power). Nevertheless, our MR analysis provided universally weak evidences to support the causal effect from cataract to T2D, which implicated the little influence of the loosen P-value threshold in this study.

In summary, we provided first and robust evidences for a sex-differential causality of T2D on cataract in East Asians, and identified several candidate genes associated with the putative causality. The results of this study provided theoretical fundament for early-stage prevention of cataract in East Asian T2D patients.

## Data Availability

The European genotypic data and phenotypic data of T2D and cataract can be assessed from the UKB project (https://www.ukbiobank.ac.uk/) after application.
The East Asian GWAS summary statistics of T2D and cataract were available in the BBJ project (http://jenger.riken.jp/en/result) and AGEN database (https://blog.nus.edu.sg/agen/summary-statistics/t2d-2020/).

## Declarations

### Ethics approval

The BBJ project was approved by research ethics committees at the Institute of Medical Science, the University of Tokyo, the RIKEN Yokohama Institute, and the 12 cooperating hospitals. The UK Biobank project was approved by the North West Multi-center Research Ethics Committee. The 22 sub cohorts used in AGEN-T2D GWAS were approved by relevant institutions.

### Availability of data and material

The European genotypic data and phenotypic data of T2D and cataract can be assessed from the UKB project (https://www.ukbiobank.ac.uk/) after application.

The East Asian GWAS summary statistics of T2D and cataract were available in the BBJ project (http://jenger.riken.jp/en/result) and AGEN database (https://blog.nus.edu.sg/agen/summary-statistics/t2d-2020/).

