## Supplementary material for "Sex differences in the risk of cataract associated with type 2 diabetes: a Mendelian randomization study": supp_fig

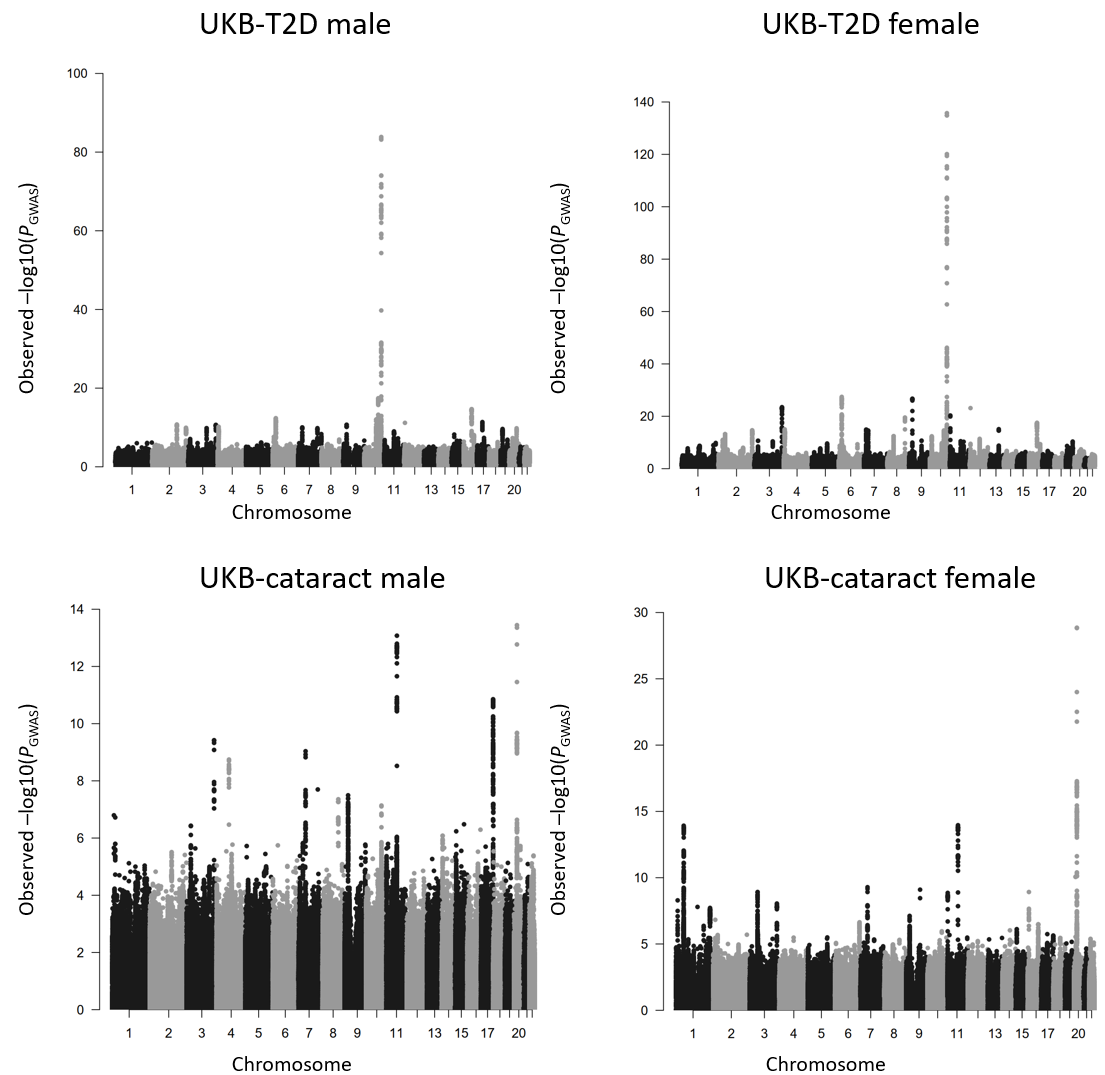

Figure S1. Manhattan plots of the sex-stratified SNP associations for T2D and cataract estimated from GWAS analysis based on the European cohort from UK Biobank.

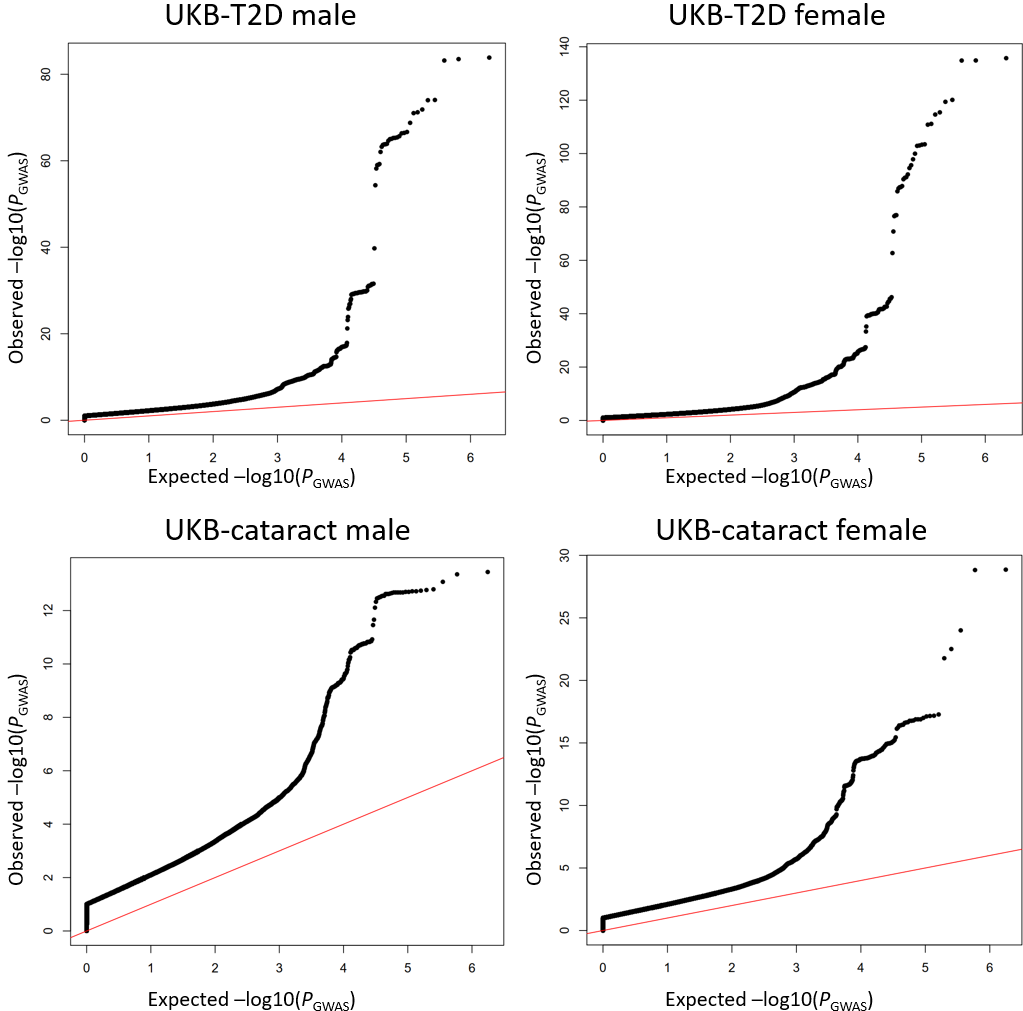

Figure S2. The quantile–quantile plots of the sex-stratified GWAS for T2D and cataract based on the UK Biobank European cohort. The red line is the diagonal line of the quantile-quantile plot.
